# Evaluation of the comparative risk of aerosol generation by tracheal intubation and extubation in the operating theatre

**DOI:** 10.1101/2020.08.24.20180067

**Authors:** Jules Brown, Florence K.A. Gregson, Andrew Shrimpton, Bryan R. Bzdek, Anthony E. Pickering, Jonathan P. Reid

## Abstract

**Background:** Transmission of SARS-CoV-2 by bioaerosols is of increasing concern. The enhanced levels of personal protective equipment (PPE) and preventative measures to attenuate viral transmission during aerosol generating procedures (AGPs) are having a huge impact on healthcare provision. There is no quantitative evidence on the number and size of airborne particles produced during AGPs to inform risk assessments.

**Methods:** Real-time, high-resolution environmental monitoring was conducted in ultraclean ventilation operating theatres. Continuous sampling with an optical particle sizer allowed characterization of aerosol generation within the airway management zone during endotracheal intubation and extubation for urgent orthopaedic trauma or neuro-surgery.

**Results:** Aerosol monitoring showed a very low background particle count allowing resolution of the transient airborne particle plume produced by reference volitional coughs (maximum concentration, 1,690±140 particles.L^-1^,n=38). By comparison, endotracheal intubation including mask ventilation produced negligible quantities of aerosolized particles (maximum concentration, 80±10 L^-1^,n=14, P<0·001 vs cough). Extubation, particularly when the patient coughed, produced a detectable aerosol plume but with a smaller number of particles (<25%) than a volitional cough.

**Conclusions:** Using a volitional cough as a reference we have been able to produce a relative risk ranking for endotracheal intubation and extubation as potential AGPs. The study does not support the assignation of endotracheal intubation by direct laryngoscopy with manual ventilation as an AGP. Extubation does generate aerosols, particularly if the patient coughs, but these are weaker than a standard reference. These findings indicate the need for a reappraisal of guidance on PPE for AGPs.

## Introduction

The SARS-CoV-2 virus and associated COVID-19 pandemic have had an unprecedented impact on global health and the world economy. Drastic interventions to limit transmission have been introduced worldwide such as physical distancing and the use of personal protective equipment (PPE). These strategies are based in part on existing knowledge from similar respiratory pathogens such as SARS-CoV-1.

Respiratory secretions have a high SARS-CoV-2 viral load and are believed to be the main source for person-to-person transmission.^1^ Coughing and sneezing atomize respiratory secretions into particles with different aerodynamic properties according to size; particles greater than 20µm in diameter are conventionally defined as droplets and tend to follow a ballistic trajectory. These droplets can either directly infect a susceptible individual within close proximity (<1m) or may settle on nearby surfaces (fomites) where viable virus can exist for up to 72 hours on surfaces.^2^ Droplet and contact transmission are considered the predominant modes of transmission of SARS-CoV-2, providing the rationale for physical distancing and hand hygiene to prevent spread.

Concern is growing about the role aerosolized viral transmission plays in COVID-19.^3,4^ Aerosolized particles (<5µm in diameter) may transmit infection by transiting the full extent of the respiratory tract to be deposited in the alveoli. The importance of airborne SARS-CoV-2 transmission relative to the droplet and contact modes remains unclear and the optimum methods of preventing bioaerosol transmission are debated.^5–8^ To minimize airborne transmission of SARS-CoV-2 to healthcare workers, specific patient care activities have been designated as Aerosol Generating Procedures (AGPs) with recommended levels of enhanced PPE. The evidence for this designation is largely based on retrospective cohort and case-control studies of transmission during the SARS pandemic; a systematic review of these studies categorized the quality of available evidence as ‘very low’ and not suitable for extrapolation to other settings.^9,10^ There is no quantitative evidence of increased aerosol generation from AGPs. Aerosol sampling in the vicinity of patients with H1N1 influenza during AGPs did not clearly demonstrate an increased risk above background of detecting virus RNA in the air (including data from intubations).^11^

Tracheal intubation and extubation, manual ventilation and respiratory tract suctioning are AGPs undertaken regularly by anaesthetists in operating theatres, exposing healthcare workers to potential airborne transmission of SARS-CoV-2.^9,12,13^ The World Health Organization has and many other governmental organization including Public Health England have recommended that such AGPs are performed in enhanced PPE consisting of a respirator style facemask, fluid resistant gown, long gloves and a face shield or visor.^13^ The evidence base for these guidelines is weak and the magnitude of risk for each AGP is unknown.^12^ There is a complete absence of evidence regarding the quantity of aerosol produced by AGPs.

Wearing enhanced levels of PPE has consequences for the conduct of clinical practice. The use of PPE for AGPs increases stress, anxiety and discomfort of healthcare workers, poses challenges for teamworking, impairs communication and these collectively have the potential to increase the risk of clinical errors. Additionally, methods introduced to mitigate the risks posed by bioaerosols have reduced operating theatre turnover, decreased hospital productivity and increased waiting times for urgent, elective and cancer surgery. A further important consideration relates to the cost and limited supply of PPE which has to be targeted to the healthcare workers at highest risk of viral infection.

The enhanced PPE recommendations are based on the assumption that AGPs pose a higher risk of aerosolized viral transmission than exposure to a patient who is coughing. There is a comparatively large evidence base around aerosolized particles generated by coughing with sizes ranging from visible droplets to submicron particles.^14–16^ When AGPs are placed in the context of a cough it is worth reflecting that in the case of intubation, the objective of the anaesthetic team is to avoid the generation of a cough - a sign of inadequate depth of anaesthesia and / or neuromuscular blockade. Given the uncertain balance of potential risks and benefits associated with the protective strategies put in place to limit viral transmission, it is important to quantitatively assess the degree to which individual AGPs generate aerosolized particles.

We aimed to quantitate particle emission in real-time during endotracheal intubation and extubation, using particle analysis instruments in a working surgical operating theatre environment. We used a volitional cough a reference comparison.

## Methods

An environmental monitoring study was conducted to quantitate the airborne particle size distribution and particle number concentration produced by AGPs in two operating theatres in a UK teaching hospital (North Bristol NHS Trust). Ethical approval for the study was given by the Faculty of Life Science and Science Research Ethics Committee at University of Bristol (ref: 105203). As this was an observational study, the anaesthetic and theatre team undertook their normal practice during airway management.

The operating theatres have an ultraclean, laminar flow ventilation system (EXFLOW 32, Howorth Air Technology) with a HEPA filtered air supply rate of 1200m ^3^.s^-1^. Temperature in theatres was set to 20°C and humidity between 40-60%. This ventilation system has a canopy, which is the ‘clean zone’ where surgical procedures are performed; the air circulation velocity is 0·2 m.s^-1^ at 1 m above the floor in the canopy. All aerosol recordings were performed in the ‘clean zone’ canopy.

A lightweight, portable Optical Particle Sizer (OPS, TSI Incorporated, model 3330) was used which samples air at 1 L min^-1^ and detects particles by laser optical scattering. The OPS reports the particle number concentration and size distribution within the optical diameter range 300 nm to 10 µm with a time resolution of 1s.

A sampling funnel was 3D printed (RAISE3D Pro2 Printer, 3DGBIRE) with a maximal diameter of 150 mm, cone height of 90 mm with a 10 mm exit port. A conductive silicone sampling tube with a length of 2 m and internal diameter 4·8 mm (3001788, TSI) was connected to the OPS to sample particles in the ‘*airway management zone*’ - the region between the patient and healthcare worker performing the AGP.

In the ultraclean theatre environment, it was possible to reliably detect a standard volitional cough (by JB) at a distance of 0·5 m between the participant and the sampling funnel, reporting a size distribution in keeping with previous aerosol studies.^17^ Sampling at 0·5 m represents a typical distance from the face of healthcare workers involved in airway management to the patient’s mouth. Nonetheless, the sampling funnel was handheld to ensure it could be quickly removed from the airway management zone in case of clinical need (this was not needed in the course of the study). All healthcare workers, and members of the investigating team, wore enhanced PPE for AGPs.

All anaesthetic inductions followed a conventional sequence with pre-oxygenation, intravenous induction by administration of anaesthetic and muscle relaxant, manual ventilation, direct laryngoscopy and intubation of the trachea followed by inflation of the endotracheal tube cuff which was the reference end point of intubation. This whole sequence typically lasted 3-4 minutes and a five-minute period before cuff up was analysed.

For extubation, the level of anaesthesia was lightened, spontaneous breathing allowed to recommence, the oropharynx suctioned before the endotracheal tube cuff was deflated, the patient extubated, and the airway patency ensured as necessary with co-application of an anaesthetic facemask before application of a Hudson mask. The reference event for extubation was ETT cuff deflation (releasing the seal on the airway) and three minutes before and up to 2 minutes after cuff down was recorded.

Airway management events were recorded contemporaneously using a time stamp application (Emerald Sequoia LLC):

### Analysis

Data were exported from the TSI optical particle sizer, processed in the TSI Aerosol Instrument Manager software, and analysed in Origin Pro (Originlab) and Prism v8 (Graphpad). Comparisons were made between aerosol generating events with unpaired t-tests with significance level at P<0·05. All data are presented as mean±SEM or ±95%CI unless otherwise indicated.

## Results

Environmental aerosol monitoring was conducted over a 3-week period during operating lists for orthopaedic trauma and neurosurgical emergencies. Recordings within the airway management zone were performed during 7 all-day theatre sessions across 3 theatres with 19 intubations and 14 extubations. The conduct of anaesthesia was left at the discretion of the anaesthetist (who ranged in experience from junior trainee to senior consultant).

Initial environmental aerosol monitoring recordings showed the ultraclean, laminar flow ventilation and air filtration system produced a very low background of aerosol particles (averaging 0·4 L^-1^ when the theatre is empty, and 3·4 L^-1^ when the theatre is in use but no AGPs are in progress). By comparison background recordings in a nearby non-laminar flow operating theatre showed aerosol particle counts that were ~4 orders of magnitude greater (15×10^3^ L^-1^).

Standard volitional coughs showed a characteristic profile with a rapid and transient jet of expectorated particles. The large majority of the particles were <1µm diameter with a concentration ~300x background when sampled at 0·5m distance. The peak aerosol concentration, 1,310±150 particles L^-1^ (n=38 coughs), typically occurred 2 seconds after the cough was registered with an average total of 134±13 particles summed over 12 seconds (Figure 1A). The spike in aerosol particle count decayed back to baseline with a time constant of ~2·7 seconds. Of note, although we conducted our monitoring in an ultraclean ventilation canopy, the cessation of flow of the ventilation system (0·2 m.s^-1^) did not alter the magnitude of the particle plume (On: 164±27 vs Off: 153±27 particles, ns, n=9 per group).

**Figure 1.**
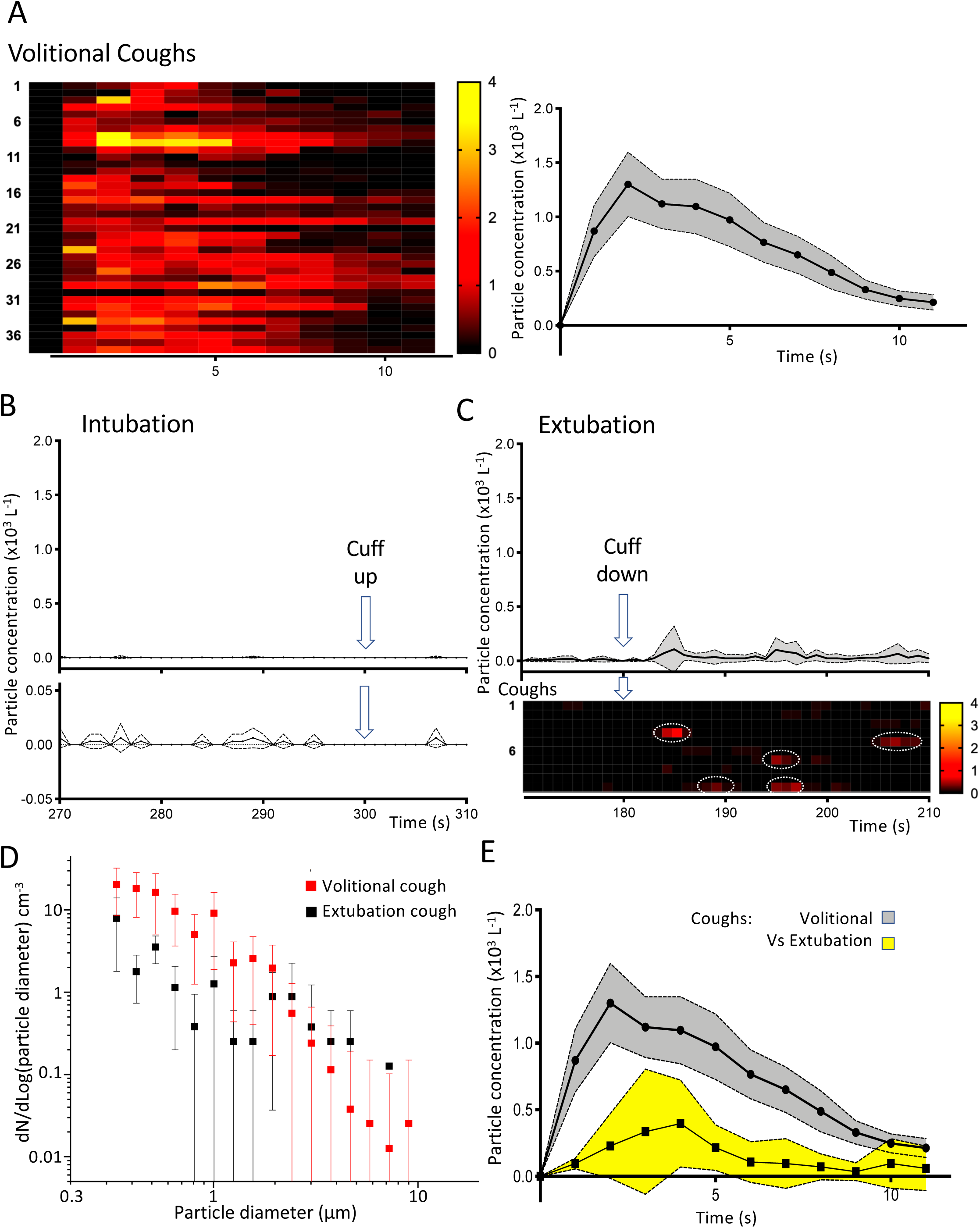
Aerosol measurement during intubation and extubation in operating theatre environment A. Temporal profile of aerosol generation from volitional coughs. Individual recordings (n=38) represented on heat map showing the total number particle concentration over time. Average time course plotted (mean±95%CI) showing a peak after 2 seconds and a decay back to baseline within 15 seconds. B. Profile of the total number concentration of aerosol detected during the critical phase of intubation (arrow at 300s marks completion of intubation with cuff up). When plotted on the same scale as the cough (B) then this looks like a flat line and when shown on a ten-fold expanded scale below it can be seen that it is not significantly different to baseline (mean±95%CI). C. Extubation recordings from each patient (n=10) plotted as the average and individually as rows on heat map of number concentration of particles (lower, on same scale as B) which showed sporadic aerosol events (red, ringed) after cuff deflation set on a low baseline level of particles. The average concentration of aerosol was low overall (shown above as mean±95%CI). D. The extubation cough events (n=5) had a similar aerosol particle size distribution to volitional coughs with a predominance of diameters <1μm (mean±SD). E. The extubation coughs were of a smaller magnitude than the volitional coughs (particle number concentration profile shown overlaid, mean±95%CI).

Monitoring during the entire period of intubation showed very little increase in aerosol generation above background levels within the airway management zone. The average number of particles detected in a 5 minute period before cuff inflation was 7±2 (n=14), compared to a background in the empty theatre of ~2 particles per 5 minute period (Figure 1B). An equivalent series of measurements during intubation in the absence of laminar flow produced a similar low particle count (6±1, n=5, ns). The average concentration of particles recorded during the intubation period (0·8 L^-1^) was around three orders of magnitude lower than the average level recorded during reference coughs (750 particles L^-1^). These recording periods always included manual mask ventilation, as well as episodes of airway suction, sometimes also requiring several attempts at laryngoscopy/intubation.

Extubation produced a relatively low average concentration of aerosolized particles (21±6 L^-1^, n=10, Figure 1C) which was ~35 fold lower than that seen during a volitional cough (750 particles L^-1^). However, given that monitoring during the extubation period was surveying over 5 minutes rather than 12 seconds this translated to a similar total number of particles (100±28 (n=10) particles over the period of extubation compared to 134±13 with a single cough, ns). It was noted that several of the extubations included a cough (often just after cuff deflation) that was timestamped and detected as an aerosol generating event (n=4/10, Figure 1C). These extubation coughs produced a similar size distribution of aerosolized particles to the standard reference coughs (Figure 1D) although they were always smaller in magnitude (33±4 particles per cough – n=5) equating to less than 25% of the particles produced by an average volitional cough (Figure 1E).

## Discussion

By conducting aerosol sampling in an ultraclean theatre environment during routine clinical practice we have been able to produce a unique, quantitative dataset documenting the aerosols produced by volitional coughs compared to common anaesthetic AGPs. These findings demonstrate the process of endotracheal intubation is associated with a very low risk of aerosol generation. Even when considering the total period of anaesthetic induction including manual ventilation and airway instrumentation (~5 minutes), the total particle count overall is still less than 5% of that associated with a single volitional cough. The conduct of a standard anaesthetic induction is designed to obtund airway reflexes and is accompanied by a muscle relaxant to ensure that the anaesthetized patient can neither breathe nor co-ordinate their upper airway musculature (vocal cords). This choreographed sequence makes the chance of an active respiratory event like a cough remote. The fact that we could only detect very low levels of aerosol indicates that slower gas movements, such as during manual ventilation or indeed with expiration due to passive elastic lung recoil when the airway is opened for laryngoscopy, generates barely detectable levels of airborne particles. It is also worth noting that these intubations reflected typical clinical practice by anaesthetists with a range of experience and were not all straightforward, providing further reassurance regarding the low level of aerosol generation.

A more nuanced picture is seen with extubation where although the overall concentration of aerosol was less than a single cough nonetheless the cumulative number of airborne particles (over a 5 minute period) was of the same order of magnitude to that seen with a single volitional cough. Indeed, a cough event was noted in 50% of extubations and this was detected as an aerosol plume. These extubation coughs were weaker than volitional coughs (~25%) and transient with aerosol only detectable for 5-10 seconds. A recognized risk during emergence from anaesthesia is that the upper airway reflexes may be obtunded by residual anaesthesia or muscle relaxant placing the patient at risk of aspiration. Hence a cough is often taken as a positive sign as it signals the return of these reflexes enabling a patient to clear secretions. The risk of aerosolized particles could be reduced by the expedient of positioning by the anaesthetist behind the patient’s head and thus out of the direct stream of any potential cough plume (as is normal practice). Coughing on extubation can also potentially be avoided by modifying the anaesthetic technique in higher risk patients.

Our results for intubation are at odds with previous retrospective evidence which was used to designate intubation an AGP^9,10^. These studies found an association between acquiring SARS and being in the room during intubation but without measuring any aerosol generation. We postulate that other mechanisms of transmission, such as direct exposure to respiratory secretions or fomites, could account for spread rather than aerosolized virus. Extubation in this study was often associated with coughs that generate a detectable aerosol, but these were weaker than a volitional cough. On this basis it seems hard to justify its designation as a high risk AGP, nonetheless appropriate precautionary measures are needed, especially in higher risk cases such as COVID positive patients. Given that a fluid resistant surgical mask is deemed an appropriate level of PPE for close exposure to a confirmed COVID-19 positive patient in current UK guidelines^13^, then use of respirator masks does not seem necessary for intubation and manual ventilation which are currently defined as AGPs and should be considered (rather than mandated) as part of a risk mitigation strategy for extubation of higher risk patients.

In conclusion this study indicates that the process of endotracheal intubation produces a barely recordable increase in aerosol and should not be defined as an aerosol generating procedure. When a patient coughs during endotracheal extubation, a measurable particle plume is produced but the aerosol is smaller than a volitional cough. These results should help inform future PPE guidelines, potentially reducing unnecessary usage of respirator type face masks whilst saving stocks for verified high risk AGPs.

## Data Availability

The data in the manuscript will be shared with any interested party on reasonable request to the corresponding author.

## Author contributions

Jules Brown - Literature search, Study design, Data collection, Data analysis, Data interpretation, Writing.

Florence Gregson - Study design, Data collection, Data analysis, Data interpretation, Figure generation, Writing.

Andrew Shrimpton - Literature search, Study design, Data collection, Data interpretation, Writing Bryan Bzdek - Study design, Data interpretation, Writing.

Anthony Pickering - Literature search, Study design, Data analysis, Data interpretation, Figure generation, Writing, Funding acquisition.

Jonathan Reid - Literature search, Study design, Data interpretation, Writing.

